# Evaluation of a Generative Medical Event Foundation Model for Predicting Post-Discharge Trajectories in Emergency Department Abdominal Pain

**DOI:** 10.64898/2026.05.18.26353199

**Authors:** Kent A. McCann, Donald S. Wright, Mark S. Iscoe, Edward R. Melnick, Lucila Ohno-Machado, Daniella Meeker, Arjun K. Venkatesh, Rohit B. Sangal, Andrew J. Loza

## Abstract

**Objective:** Emergency department (ED) risk models predict whether a patient will revisit within a fixed window, but not what the visit will entail. We evaluated whether a medical event foundation model predicts the timing, sequence, and severity of downstream care after ED discharge for abdominal pain.

**Materials and Methods:** Retrospective cohort study in the de-identified Epic Cosmos network. From 150,030 eligible adults, we analyzed a random sample of 3,000, generating 50 simulated 30-day trajectories per patient with Curiosity, a generative medical event foundation model, and compared these with gradient boosted tree baselines. Endpoints were discrimination for three outcome classes (any revisit, discharge-revisit, admit-revisit) at three horizons, 36-class trajectory accuracy, transition-level calibration, and subgroup performance.

**Results:** Curiosity’s 30-day admit-revisit AUROC was 0.83 (95% CI 0.79-0.87) versus 0.76 (0.71-0.81) for XGBoost (P=.002). Differences were significant at five of nine outcome-horizon pairs but not for discharge-revisit at any horizon. Its most likely trajectory (of 36) matched the observed in 45.9% versus 43.0% (P<.001); median absolute calibration error across 45 transitions was 1.30 percentage points. Discrimination and calibration were poorer for Hispanic or Latino, self-pay, and other-race patients.

**Discussion:** Without task-specific training, one pretrained foundation model matched or exceeded a tuned supervised baseline, with significant advantages on the primary endpoint and trajectory prediction but none for discharge-revisit. Subgroup gaps must be resolved before clinical use.

**Conclusion:** A medical event foundation model can represent post-ED-discharge revisit risk as a calibrated trajectory rather than a single probability; prospective evaluation and remediation of subgroup performance gaps remain necessary.

## Background and Significance

Medical event foundation models are generative models pretrained on longitudinal coded electronic health record (EHR) events that simulate future sequences of care (diagnoses, laboratory results, procedures, medications, encounters, and elapsed time) from a patient’s current state.[1–3] Because a single pretrained model can generate predictions for many endpoints without task-specific training, these models could support decision making across outcomes that would otherwise each require a separate supervised model, training set, and deployment pipeline. Evaluations to date, however, have emphasized broad single-task panels or disease-incidence prediction.[1,2] No published evaluation pairs per-outcome discrimination across multiple horizons with multi-step trajectory accuracy, transition-level calibration, and subgroup performance on a single clinically defined cohort, benchmarked against a supervised model comparator.

Health systems face this question operationally: foundation models trained on network-scale EHR data are becoming available within vendor ecosystems, and institutions must decide whether to deploy them, typically without the ability to retrain the model or inspect its training data. Such decisions require a transferable evaluation pattern: a clinically defined cohort, endpoints spanning discrimination, trajectory accuracy, and calibration, a supervised baseline given enough training data and tuning to represent a reasonable comparator, and a subgroup audit tied to the deployment decision. This study operationalizes that pattern for one model and one decision point.

Abdominal pain is the most common reason for emergency care in the United States (∼10 million visits annually), most visits end in discharge, and disposition decisions carry residual uncertainty about symptom course, revisit, and hospitalization.[4–9] Existing revisit-prediction research collapses the post-discharge course into binary outcomes,[10,11] and revisit risk varies across racial, ethnic, and insurance groups,[12,13] making subgroup performance directly relevant to deployment.

## Objective

We evaluated Curiosity,[3] a generative medical event foundation model, in adults discharged from the ED with abdominal pain, to determine whether simulated trajectories characterize the timing, sequence, and severity of post-discharge revisits with discrimination and calibration superior to task-specific count-based gradient-boosted tree baselines (XGBoost).

## Materials and Methods

### Study Design and Setting

We conducted a retrospective cohort study using EHR data from Epic’s Cosmos research network, a de-identified dataset of over 300 million patient records from more than 300 health systems in the United States, Canada, and the Middle East.[14,15] Curiosity is a decoder-only transformer (1 billion parameters) with a 7,105-token event vocabulary and an 8,192-token context window, pretrained autoregressively on 115 billion discrete medical events (151 billion tokens) from 118 million patients within this network.[3] Given a patient’s prior medical history, Curiosity sequentially generates future events (diagnoses, laboratory results, procedures, medication orders, encounters, and elapsed time), enabling patient-level simulation of post-discharge trajectories. It does not incorporate vital signs, clinical notes, imaging results, or medication administration records. This study was exempted from human subjects review by the Yale University Institutional Review Board.

### Study Population

From patients meeting Curiosity’s eligibility criteria for the non-training partition (age 18 January 1, 2012, with 2 encounters), we identified index ED encounters between January 1, 2022, and December 31, 2022, for which the structured chief-complaint field was coded as abdominal pain and the visit ended in discharge. For patients with multiple qualifying encounters, the first chronological encounter was retained, as repeat presenters are a clinically distinct population from patients with initial undifferentiated abdominal pain.[16] A random sample of 3,000 patients was drawn from the full eligible cohort, a size chosen to balance the computational cost of 50 simulations per patient against the precision of the primary admit-revisit comparison. Model inputs were derived directly from recorded EHR events, and no imputation was performed.

### Trajectory Simulation

For each patient, 50 independent simulations were generated from the moment of ED departure, each 512 tokens long, corresponding to approximately 390 future medical events at the corpus-average of 1.3 tokens per event.[3] This length was chosen empirically to maximize how many simulations reached 30 days post-discharge. Model parameters were not fine-tuned; all simulations used the pretrained Curiosity model with default settings.[3] Performance plateaued by 50 simulations or had minimal predicted further improvement (Figure S1).[17] To accommodate the model’s maximum input length, only the most recent 8,192 tokens from each patient’s history were used as input (see Waxler et al[3] for tokenization details). Computationally, each prediction requires autoregressive generation rather than a single forward pass: 50 simulations of 512 tokens each amount to 25,600 generated tokens per patient, executed once at the prediction point. Per-patient wall-clock time was not measured on the shared research infrastructure and would depend on production hardware.

### Outcomes

All encounters within 30 days after discharge, both observed and simulated, were classified into three categories: DC-revisit (ED revisit ending in discharge), admit-revisit (ED revisit ending in admission), and Outpatient Contact, which was further categorized into In-Person Visit, Telehealth/Message, Lab/Imaging, or Pharmacy (Table S1). Admit-revisit comprised encounter types ending in hospitalization (Table S1); observation-status and inpatient admissions were treated as a single admission category.

Encounters not mapping to any listed category were excluded from trajectory construction. Trajectories were assessed across three sequential windows (0–72 hours, 72 hours–7 days, 7–30 days), each assigned one of three outcomes: no ED revisit, DC-revisit, or admit-revisit. ED revisits were coded as terminal, so subsequent windows were not evaluated, yielding 45 transitions across 36 complete trajectories (Table S2). A patient labeled “no revisit” (shortened from “no ED revisit” in figures) at the 7–30 day window therefore had no revisit at any point in the 30-day period.

### Comparator: XGBoost Baselines

We compared performance to a conventional machine learning approach commonly used in prior ED revisit prediction studies.[10,11] As a benchmark, we trained an XGBoost classifier[18] using each patient’s EHR history represented as counts of clinical events with the same feature vocabulary used as Curiosity input. This count-based representation, standard in this literature, records event frequency but not order or timing. Unlike the Curiosity input (capped at 8,192 tokens), XGBoost feature counts were computed over each patient’s complete pre-index history. Training cohorts were drawn from the Cosmos training partition, identified with the same 2022 index-encounter eligibility criteria and with no overlap with the evaluation cohort; training and evaluation cohorts were demographically similar (Table S5). Separate models were fit for each time horizon (72 hours, 7 days, 30 days), predicting a three-class outcome: no ED revisit, DC-revisit, or admit-revisit. A fourth model was trained to predict the 36-category trajectory directly. For the learning-curve sweep, hyperparameters followed Waxler et al[3]; the final comparator models were hyperparameter-tuned as described below (Table S3). Models were evaluated on the same 3,000-patient cohort as Curiosity for direct paired comparison.

#### Baseline learning curve and tuning

To ensure the comparison was not an artifact of a data-starved or under-specified baseline, we trained the XGBoost models on nested training samples of increasing size (10,000 to 100,000 patients in 10,000-patient increments; 5 random seeds per size) with identical features and fixed hyperparameters (Table S3), and report the resulting learning curve. Each of the four models (three per-horizon classifiers and the 36-class trajectory model) was then independently hyperparameter-tuned at the observed plateau of 100,000 training patients using Bayesian optimization (Optuna[19], 40 trials per endpoint with 3-fold cross-validation; Table S3), with final models refit on the full 100,000-patient training sample as an ensemble across the same 5 random seeds. These tuned models served as the primary comparator for all head-to-head analyses.

### Statistical Analysis

#### Per-outcome discrimination

For each of three outcome classes (admit-revisit, DC-revisit, any ED revisit) at each of three post-discharge horizons (72 hours, 7 days, 30 days), we compared Curiosity and XGBoost discrimination by area under the receiver operating characteristic curve (AUROC) and area under the precision-recall curve (AUC-PR). The primary comparison was 30-day admit-revisit AUROC, on which the sample size was based; the remaining eight comparisons were secondary, with unadjusted P values interpreted as nominal. Curiosity’s predicted probability for each outcome-horizon pair was the proportion of the patient’s 50 simulations containing the target outcome by that horizon; for XGBoost, the appropriate class probability (or sum of revisit-class probabilities for any ED revisit) was used. AUROCs were compared using DeLong’s test for paired ROC curves;[20] 95% bootstrap CIs were constructed with patient as the resampling unit (Supplementary Methods).

#### Trajectory-level accuracy

Each patient’s 30-day post-discharge course was classified into one of 36 trajectory classes (Table S2). Trajectory similarity was assessed using edit distance (the number of insertions, deletions, or substitutions required to convert a predicted trajectory to the ground truth).[21] For Curiosity, each patient’s edit distance was the mean across all 50 simulated trajectories; for XGBoost, it was the expected edit distance under the 36-class trajectory model’s softmax probabilities. Per-patient values were compared between models via paired Wilcoxon signed-rank test. We compared the proportions of patients for whom the observed trajectory was Curiosity’s single most probable prediction, among its three most probable, or among its five most probable, using McNemar’s tests with Bonferroni correction for the three comparisons ( = 0.017).

#### Stepwise calibration of conditional path probabilities

For each of the 45 transitions, we computed the conditional probability of reaching each downstream event given passage through the upstream event from ground truth and simulated trajectories. Calibration accuracy was quantified as the median absolute difference between simulated and observed transition probabilities across all 45 transitions; a 95% CI was derived from a patient- and simulation-level hierarchical bootstrap.[22] Calibration metrics for XGBoost were computed similarly (Supplementary Methods).

#### Subgroup performance and calibration

To assess consistency across patient subgroups, we estimated AUROC and logistic calibration slope within demographic, acuity, and insurance subgroups for 30-day any ED revisit, the only endpoint with enough events per stratum for stable estimates. Calibration slopes were estimated on empirical-logit-transformed simulation proportions (Supplementary Methods); values below 1 indicate over-extreme risk estimates. Strata with fewer than 11 events or fewer than 11 non-events were suppressed per the Epic Cosmos minimum cell-size policy.

#### Class imbalance and recalibration

No class-imbalance resampling or reweighting was applied to either model; for low-prevalence outcomes such as admit-revisit, we report AUC-PR against the observed base rate alongside AUROC. Predictions from both models were evaluated as generated, without post hoc recalibration.

#### Trajectory visualization

We visualized 30-day trajectories using parallel Sankey diagrams for observed and simulated distributions, and decomposed outpatient engagement within each time window using UpSet plots.[23] To display trajectories for patients at the head of each risk distribution, we ranked the cohort separately by simulation-derived admit-revisit and DC-revisit probability and took the top 5% by each ranking (n = 150 per group), a descriptive display slice rather than a pre-specified analysis threshold. The same ranking procedure was applied to XGBoost predicted probabilities for comparison.

#### Reporting

This study is reported in accordance with the TRIPOD+AI statement for prediction-model studies involving machine learning;^24^ the completed checklist is provided in the Supplementary Appendix.

#### Software

Analyses were performed in Python 3.13 with pandas, numpy, scipy, statsmodels, scikit-learn, xgboost, and optuna; figures in matplotlib and plotly (full versions in Supplementary Methods). Two-sided P<.05 was considered statistically significant except as noted above in cases of multiple comparisons.

## Results

### Study Population

From 179,310 qualifying ED encounters, 150,030 unique patients were eligible after deduplication. Among eligible patients, 6.4% were Emergency Severity Index (ESI) 1–2, 89.2% ESI 3, 3.6% ESI 4–5, and 0.7% had undocumented ESI. The evaluation cohort comprised a random sample of 3,000 patients from the full eligible cohort; baseline characteristics are presented in Table 1.

**Table 1.** Characteristics and Cumulative Revisit Rates of the Evaluation Cohort. A simple random sample of 3,000 patients was drawn from 150,030 eligible patients. Descriptive statistics are presented as median [IQR] for continuous variables and n (%) for categorical variables. Emergency Severity Index (ESI) levels are collapsed (1–2 and 4–5) in accordance with the Epic Cosmos minimum cell-size policy.

|  | Overall |
| --- | --- |
| n | 3,000 |
| Age, median [IQR] | 47.0 [36.0–60.3] |
| **Revisits (cumulative)** |  |
| 72 Hour | 162 (5.4%) |
| 7 Day | 249 (8.3%) |
| 30 Day | 506 (16.9%) |
| **Sex** |  |
| Female | 1,958 (65.3%) |
| Male | 1,019 (34.0%) |
| Other/Unspecified | 23 (0.8%) |
| **Race** |  |
| White | 1,738 (57.9%) |
| Black or African American | 627 (20.9%) |
| Not Reported | 348 (11.6%) |
| Other | 287 (9.6%) |
| **Ethnicity** |  |
| Not Hispanic or Latino | 2,153 (71.8%) |
| Not Reported | 447 (14.9%) |
| Hispanic or Latino | 400 (13.3%) |
| **Acuity Level** |  |
| ESI 1–2 – Immediate/Emergent | 193 (6.4%) |
| ESI 3 – Urgent | 2,672 (89.1%) |
| ESI 4–5 – Less/Non-Urgent | 111 (3.7%) |
| Unspecified | 24 (0.8%) |

### Simulation Performance

Of the 150,000 generated simulations (3,000 patients, 50 each), 149,720 (99.81%) reached either the 30-day horizon or a terminal ED revisit and were analyzed; the remaining 280 (0.19%, affecting 4.5% of patients) were excluded. Encounter-type mapping and exclusion rates are detailed in Table S1.

### Population-Level Trajectory Distributions

Most patients followed a “no ED revisit” trajectory across all three time windows, with simulated and observed distributions in close agreement (Figure 1A). At 72 hours, 94.6% of patients had not revisited the ED (simulated: 92.2%); by 30 days, 83.1% remained without an ED revisit (simulated: 78.3%). Outpatient contact increased across windows, reaching 40.4% by 30 days (simulated: 39.0%). Figure 1B shows the composition of outpatient contact by encounter-type combination at each time window.

**Figure 1.**
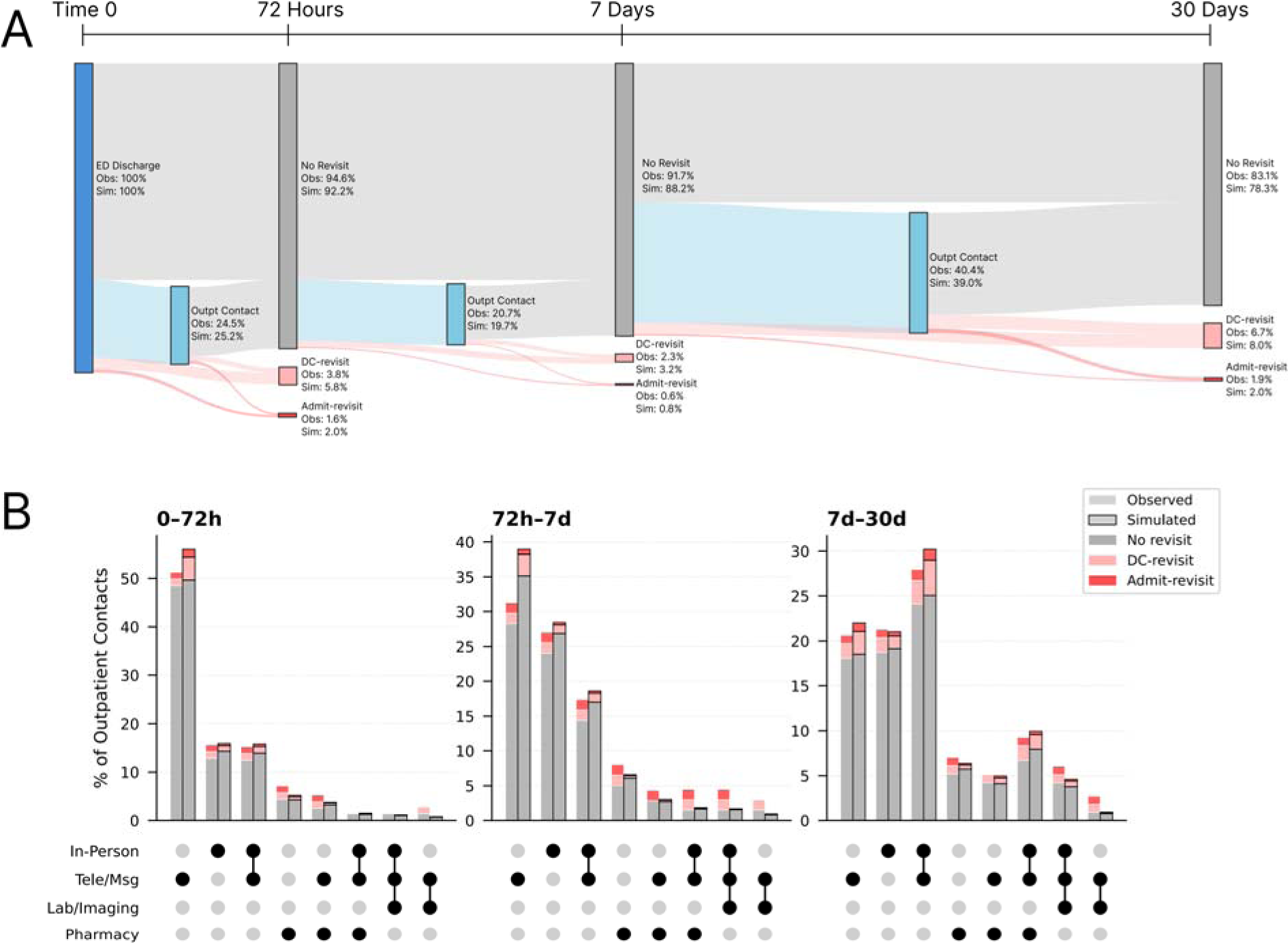
Cohort-Level Post-Discharge Trajectory Distributions. (A) Sankey diagram showing patient flow across three post-discharge time windows (0–72 hours, 72 hours–7 days, 7–30 days) for 3,000 patients with abdominal pain discharged from the ED. Link widths and node sizes represent simulated trajectory proportions (n = 150,000 simulations; 50 per patient). Node annotations show simulated versus observed percentages at each state. Outpatient Contact nodes aggregate all outpatient encounters (In-Person, Tele/Msg, Lab/Imaging, Pharmacy). (B) UpSet plots showing the frequency of specific outpatient contact combinations within each time window, comparing observed (solid) and simulated (hatched) distributions as a proportion of trajectories with any outpatient contact. Bar segments are colored by clinical outcome. Bar heights in panel B were rounded in accordance with the Epic Cosmos minimum cell-size policy.

### Per-Outcome Discrimination

Curiosity’s AUROC was higher than XGBoost’s at all nine outcome-by-horizon combinations, and significantly higher at five: all three any-revisit horizons and admit-revisit at 7 and 30 days (Table 2). Discrimination was strongest for admit-revisit, with Curiosity AUROC 0.83 (95% CI 0.79–0.87) at 30 days versus 0.76 (0.71–0.81) for XGBoost (DeLong P = .002), and 0.85 versus 0.79 at 72 hours (P = .18). For any ED revisit and DC-revisit, Curiosity AUROCs ranged from 0.70 to 0.74, with absolute differences over XGBoost of 0.01 to 0.06; the DC-revisit differences did not reach statistical significance. AUC-PR, which is sensitive to outcome prevalence, showed the same numerical ordering: at 30 days, Curiosity admit-revisit AUC-PR was 0.37 (95% CI 0.29–0.46) against a 4.1% cohort prevalence, approximately 1.7-fold higher than XGBoost AUC-PR of 0.22 (95% CI 0.15–0.29).

**Table 2.**
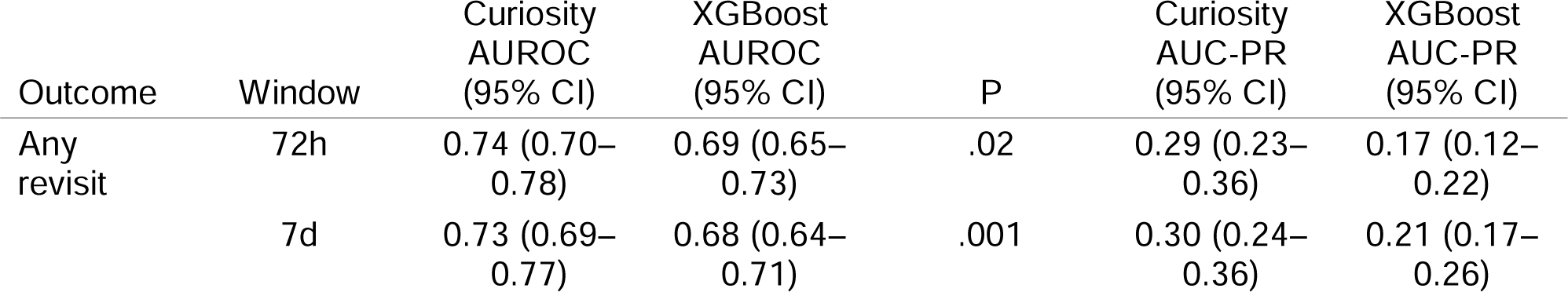
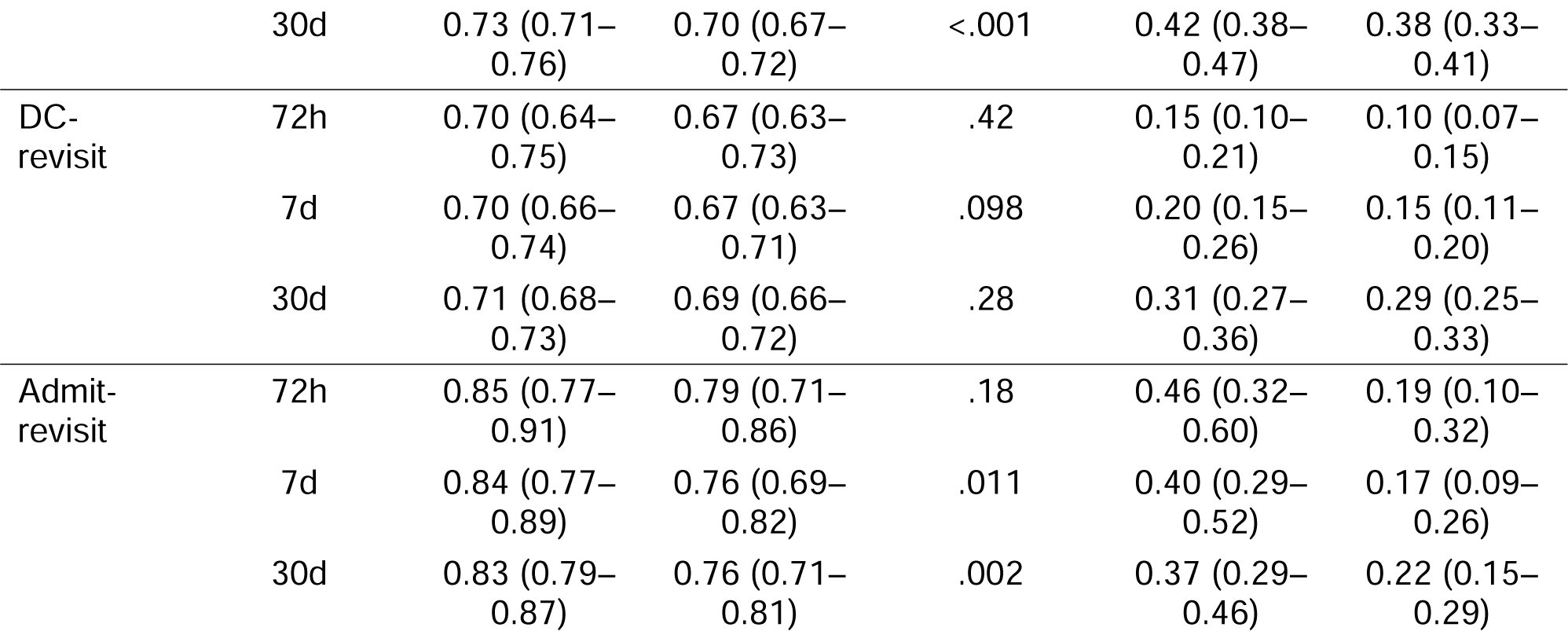
Per-Outcome Discrimination Across Post-Discharge Time Horizons. Area under the receiver operating characteristic curve (AUROC) and area under the precision-recall curve (AUC-PR), each with 95% bootstrap confidence intervals, for Curiosity and XGBoost across three outcome classes (any ED revisit, DC-revisit, admit-revisit) and three post-discharge time windows (72 hours, 7 days, 30 days). Curiosity predicted probabilities were derived from the proportion of each patient’s 50 simulations containing the target outcome by the specified horizon. XGBoost predicted probabilities were derived from per-horizon three-class classifiers (no ED revisit / DC-revisit / admit-revisit), with the appropriate class probability or class-probability sum used for each outcome. Bootstrap confidence intervals were constructed with patient as the resampling unit. P values are from DeLong’s test for paired ROC curves; AUC-PR is reported without a formal paired test. XGBoost values are from models trained on 100,000 patients, at the plateau of the training-size learning curve (Figure S2), with per-endpoint hyperparameter tuning (Table S3). Outcome base rates in the evaluation cohort, which set the AUC-PR baseline under random ranking, are 16.9% for any ED revisit by 30 days, 12.8% for DC-revisit, and 4.1% for admit-revisit (categories rounded independently). Individual-level calibration metrics (calibration slope, calibration-in-the-large, and integrated calibration index) for all nine endpoints appear in Table S4.

### Baseline Learning Curve

Under the fixed development hyperparameters, XGBoost discrimination for the primary endpoint (30-day admit-revisit) rose from AUROC 0.71 at 10,000 training patients to a plateau by approximately 60,000 patients (0.75–0.76), beyond which mean AUROC across five seeds changed by <0.01 per doubling (0.749 at 50,000 to 0.756 at 100,000; Figure S2). The 36-class trajectory model showed a noisier curve, rising through 80,000 patients, with mean macro one-vs-rest AUROC within 0.012 across the 80,000–100,000 range. At 100,000 patients each model was independently tuned; the tuned models (primary-endpoint AUROC 0.76; Table 2) served as the comparator for all head-to-head analyses.

### Trajectory-Level Accuracy

Of 36 possible trajectory classes, Curiosity’s most likely predicted trajectory matched the observed trajectory for 45.9% of patients, compared with 43.0% for XGBoost (McNemar’s P<.001). For reference, the modal observed trajectory (no outpatient contact and no ED revisit in any window) occurred in 39.8% of patients, the accuracy that a naive baseline always predicting the modal class would achieve. The correct trajectory appeared among Curiosity’s three most probable predictions for 69.3% of patients versus 65.5% for XGBoost (McNemar’s P<.001), and among the five most probable for 80.4% versus 79.2% (P=.08). Trajectory edit distance also favored Curiosity (median 1.28 vs 1.38; median paired difference 0.15, 95% CI 0.12–0.18; paired Wilcoxon P<.001), with Curiosity producing the closer trajectory in 1,819 of 3,000 patients (60.6%).

### Calibration

At the individual level, Curiosity’s calibration slopes were near 1 for admit-revisit at all horizons (1.03–1.11) and ranged from 0.72 to 0.83 for DC-revisit and any-revisit endpoints, while calibration-in-the-large was negative for all nine (−0.84 to −0.42), indicating well-scaled risk differentiation with over-prediction of absolute risk on average (Table S4). The tuned XGBoost models were similarly well calibrated at the individual level, with slopes modestly above 1 (1.06–1.31) and calibration-in-the-large near 0 (−0.09 to 0.03, all 95% CIs crossing 0; Table S4). At the trajectory level, across the 45 transitions spanning the three time windows, the median absolute difference between simulated and observed conditional path probabilities was 1.30 percentage points (95% CI 0.32–2.49 pp; Figure 2).

**Figure 2.**
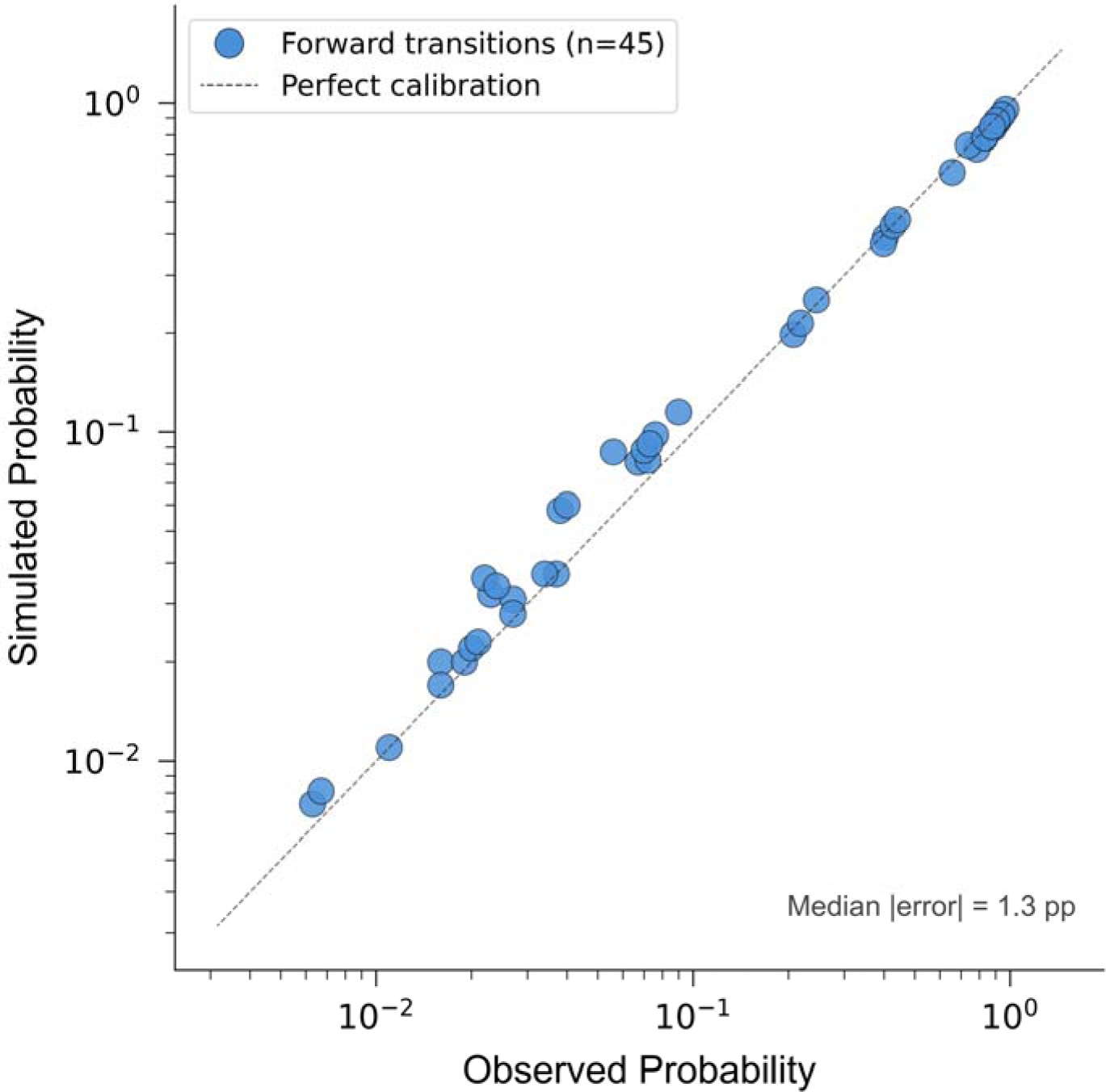
Stepwise Calibration of Simulated Trajectory Probabilities. Log-log scatter of simulated (y-axis) against observed (x-axis) conditional path probabilities for each of the 45 transitions that compose the three-window outcome paths. Each point represents one transition (e.g., ED Discharge Outpatient Contact 7d, or No Revisit 7d Admit-Revisit 30d); the dashed diagonal indicates perfect calibration. Across all 45 transitions, the median absolute difference between simulated and observed probabilities was 1.30 percentage points (95% bootstrap CI 0.32–2.49 pp).

### Trajectory Differentiation Among Top-Risk Patients

Trajectories differed between Curiosity’s high admit-revisit-risk and high discharge-revisit-risk groups (Figure 3), which were largely non-overlapping (13 patients in both). Observed admit-revisit rates were 35.3% in the high admit-revisit-risk group versus 7.3% in the high discharge-revisit-risk group (relative risk [RR] 4.82); DC-revisit rates were 47.3% in the high discharge-revisit-risk group versus 16.0% in the high admit-revisit-risk group (RR 2.96). Relative to the full cohort (admit-revisit 4.1%, DC-revisit 12.8%), these correspond to RRs of 8.62 and 3.71 (Figures 3A and 3B). Applying the same top-5% selection to predicted probabilities from the XGBoost three-class 30-day model identified an enriched admit-risk group (cohort-referenced RRs 6.02 for admit-revisit and 2.72 for DC-revisit) but showed weaker separation between the two risk subtypes: group overlap was 18.7% (vs 8.7% for Curiosity), and the cross-group DC-revisit RR was 1.33 (vs 2.96 for Curiosity).

**Figure 3.**
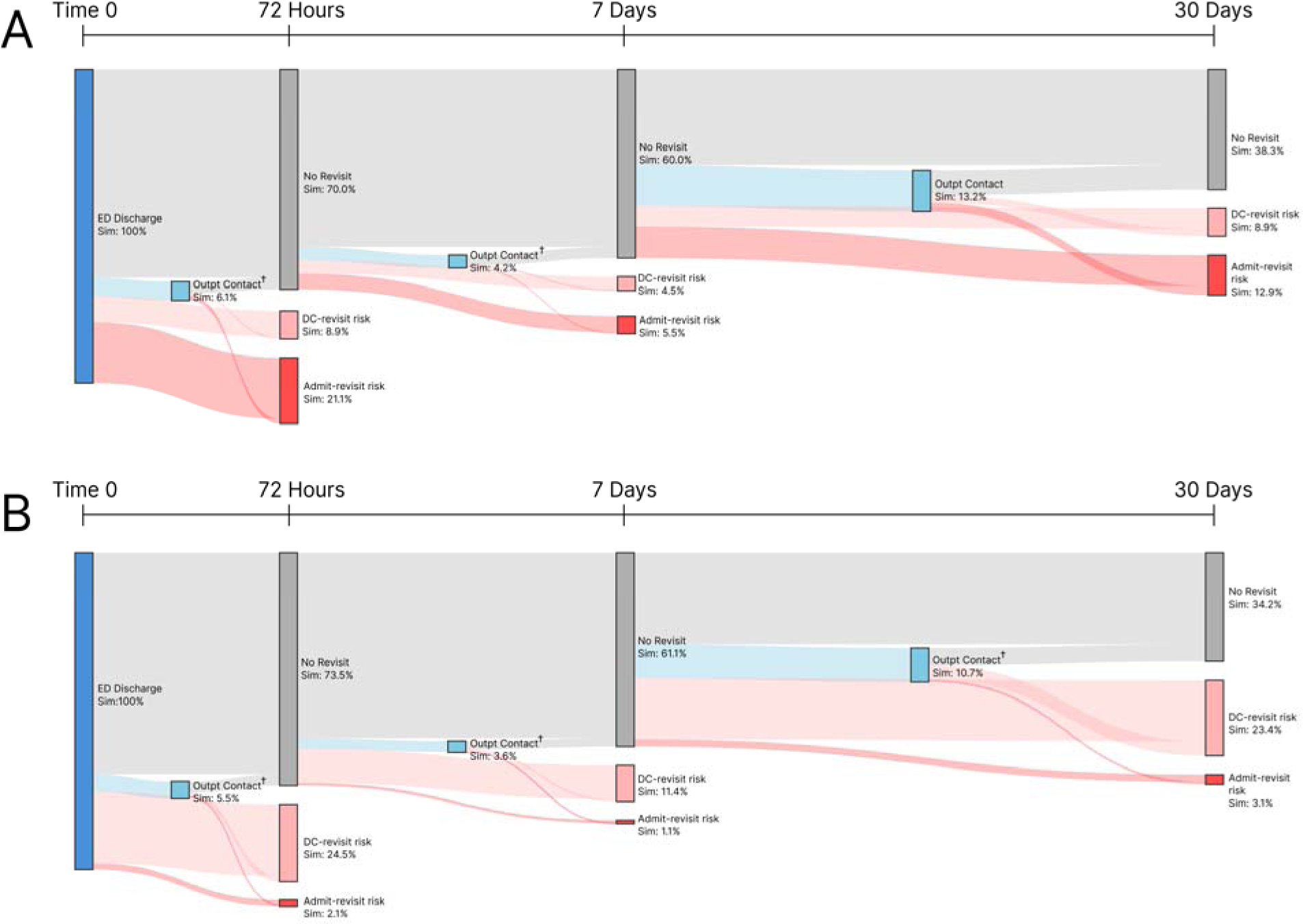
Simulated Post-Discharge Trajectories in the High Admit-Revisit-Risk and High Discharge-Revisit-Risk Groups. (A) Sankey diagram showing simulated post-discharge trajectories for the high admit-revisit-risk group, defined as the 150 patients (top 5% of the evaluation cohort) with the highest Curiosity simulation-derived probability of admit-revisit by 30 days, computed as the fraction of each patient’s 50 simulations ending in an admit-revisit. (B) Sankey diagram showing simulated post-discharge trajectories for the high discharge-revisit-risk group, defined as the 150 patients (top 5%) with the highest Curiosity simulation-derived probability of DC-revisit by 30 days. Panels A and B use the same node schema as Figure 1A; link widths represent simulated trajectory proportions within each group. A total of 13 patients (8.7% of each group) met the top-5% cutoff for both groups and appear in both panels rather than being excluded. †Denotes nodes where link sizes were rounded in accordance with the Epic Cosmos minimum cell-size policy.

### Subgroup Performance

To assess consistency across patient subgroups, we examined discrimination and calibration for 30-day any ED revisit, the only endpoint with enough events per stratum to support stable subgroup estimates; admit-revisit (123 events overall) produced subgroup cells below the reportable threshold. Overall any-revisit AUROC was 0.73 (95% CI 0.71–0.76). Discrimination was similar for women and men (0.72 and 0.75) and increased modestly with age (0.70 for ages 18–44 to 0.78 for 65 and older; Table 3). Performance was lower among patients of Hispanic or Latino ethnicity (AUROC 0.64, 95% CI 0.56–0.71) than non-Hispanic patients (0.75, 95% CI 0.72–0.78), among self-pay patients (0.58, 95% CI 0.49–0.69), and in the smaller other-race stratum (0.56, 95% CI 0.42–0.70). Calibration slopes ranged from 0.29 to 1.31 across subgroups and were most attenuated in the same strata showing lower discrimination (other race 0.29, self-pay 0.42, Hispanic or Latino 0.48). Strata with fewer than 11 events or fewer than 11 non-events were suppressed in accordance with the Epic Cosmos minimum cell-size policy.

**Table 3.**
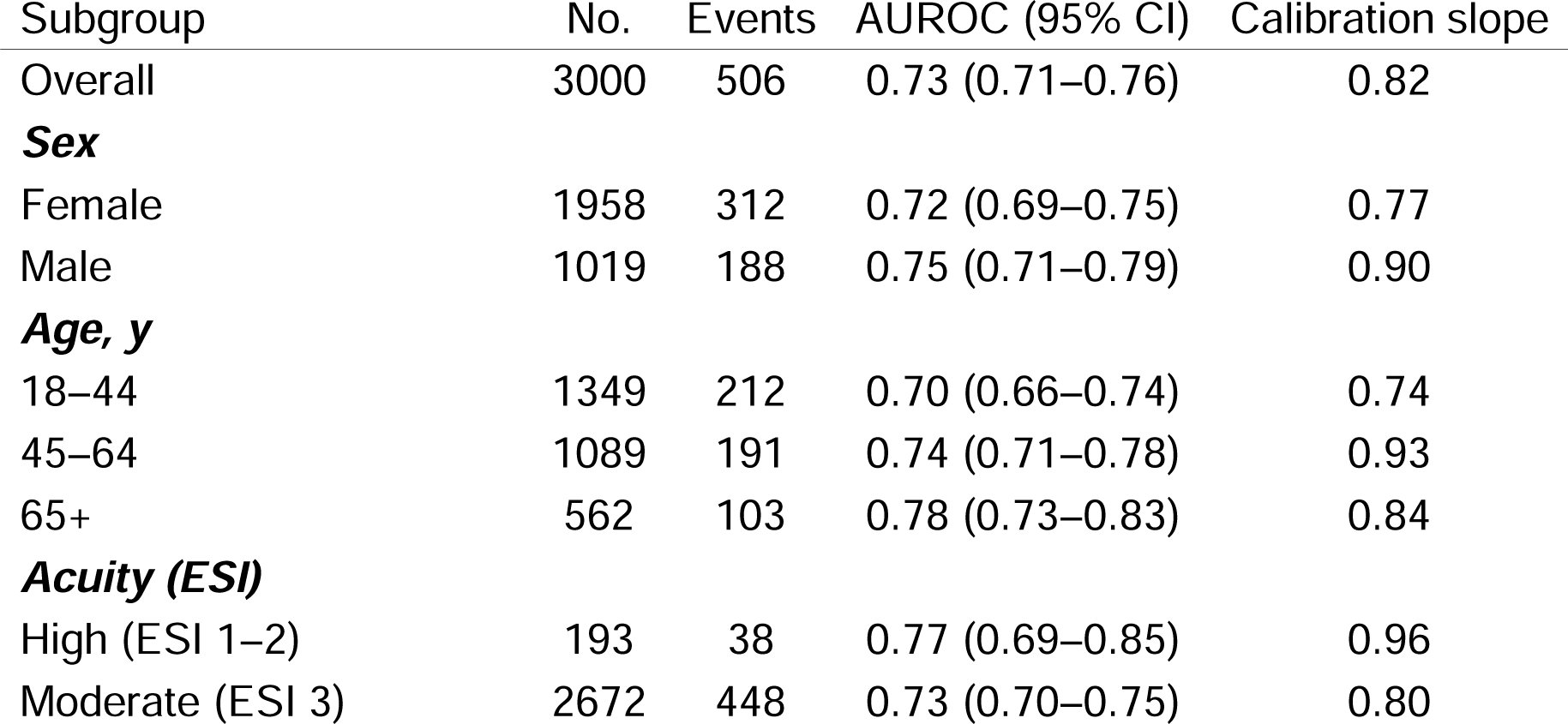
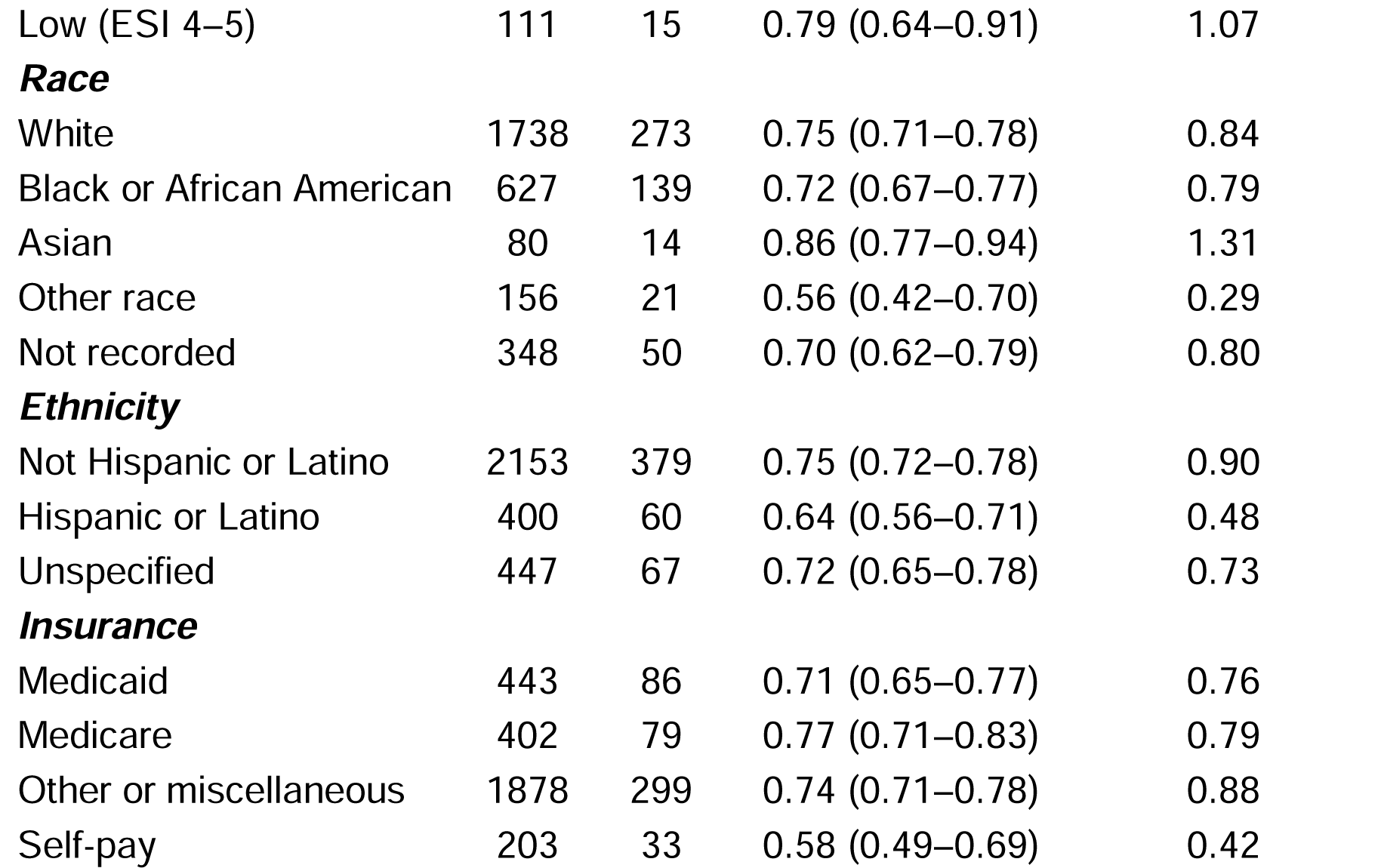
Subgroup Performance for 30-Day Any ED Revisit. Area under the receiver operating characteristic curve (AUROC) with 95% bootstrap confidence interval and logistic calibration slope (1.0 indicates ideal calibration) for 30-day any ED revisit within demographic and acuity subgroups of the 3,000-patient evaluation cohort. The any-revisit endpoint was used for subgroup analysis because admit-revisit (123 events cohort-wide) produced cells below the reportable size in most strata; the primary admit-revisit discrimination result appears in Table 2. Subgroups with fewer than 11 events or fewer than 11 non-events were suppressed per the Epic Cosmos minimum cell-size policy and are omitted (sex: ambiguous; acuity: unknown; race: American Indian or Alaska Native, Native Hawaiian or Other Pacific Islander; insurance: not applicable, unspecified). Acuity bands collapse Emergency Severity Index (ESI) levels (High, ESI 1–2; Moderate, ESI 3; Low, ESI 4–5). Calibration slopes were estimated on empirical-logit-transformed simulation proportions (Supplementary Methods); values below 1 indicate over-extreme risk estimates.

## Discussion

### Principal Findings

In this multi-system retrospective cohort comparison of foundation-model and supervised machine-learning predictions of post-ED-discharge trajectories, a generative EHR model predicted clinically recognizable post-discharge trajectories, capturing whether patients returned, when they returned, and whether the revisit ended in discharge or admission. Without task-specific fine-tuning, Curiosity outperformed the tuned XGBoost baseline on the primary endpoint and four of eight secondary comparisons, with the largest absolute gains for admit-revisit (AUC-PR 0.37 vs 0.22, approximately 1.7-fold higher against a 4.1% base rate). Trajectory-level evidence supported these results: the model matched the exact observed trajectory out of 36 classes for 45.9% of patients (versus 39.8% for a naive modal-class baseline and 43.0% for XGBoost), produced predicted trajectories closer to observed than XGBoost in 60.6% by edit distance, and tracked observed conditional transitions (median absolute error1.30 percentage points across 45 transitions).

### Beyond Binary Prediction

Beyond predicting whether a revisit occurs, a medical event foundation model distinguished what kind of revisit it was likely to be. Ranking patients separately by predicted admit-revisit and DC-revisit probability identified two largely non-overlapping groups at the heads of their respective risk distributions, each carrying an elevated rate of the corresponding revisit type. Figure 3 illustrates this at a descriptive top-5% slice: among the 150 highest-ranked by admit-risk, 35.3% subsequently returned requiring admission (vs 4.1% in the full cohort), and among the 150 highest-ranked by DC-risk, 47.3% returned with discharge (vs 12.8%).

Prior evaluations of generative medical event foundation models have demonstrated capability across broad single-task panels or disease-incidence predictions.[1–3] None addresses the question examined here: per-outcome discrimination across multiple horizons in a clinically defined cohort, paired with multi-step trajectory prediction and calibration. On the side of purpose-built ED-revisit classifiers, a 2024 scoping review reported AUROCs spanning 0.70 to 0.85 across 14 studies,[10] with recent examples spanning AUROC 0.705 to 0.845.[25–27] Curiosity’s any-revisit AUROCs of 0.73–0.74 fall within this range without task-specific training; for admit-revisit, not separately reported in this literature, its AUROC of 0.83 at 30 days sits at the upper end of the range for undifferentiated revisit. We tested a single family of supervised models; other well-tuned approaches might achieve higher performance than the baseline reported here. The case for a generative model does not rest on beating a specialized classifier at a single endpoint: one pretrained model delivered calibrated predictions across outcomes and horizons that would otherwise each require a separate model, training set, and deployment pipeline, shifting the bottleneck toward identifying which predictions merit clinical use. Other institutions facing the same decision about a proprietary foundation model they cannot retrain or inspect can reuse the evaluation pattern applied here: a clinically defined cohort, endpoints spanning discrimination, trajectory accuracy, and calibration, a supervised baseline, and a subgroup audit.

### Integration Considerations

Any clinical use of these predictions is untested and would be a hypothesis for implementation research. Two practical points bear on eventual integration. First, the model runs on data the EHR already contains: because Curiosity was trained within the same network, the coded event history it requires is present in the production record as-is, with no separate feature-engineering or terminology-mapping step. This is an advantage over externally developed models, though one that ties the model to its vendor ecosystem. Second, generating trajectories is computationally intensive and would be done once per patient at the decision point, making the predictions better suited to workflows that do not require immediate real-time inference. Finally, unlike a conventional risk score, each prediction is backed by simulated care sequences that can be read and reviewed. How best to summarize them for clinicians at the point of care is itself a question that would need dedicated evaluation before use.

### Subgroup Performance and Readiness for Clinical Use

Subgroup analyses revealed performance gaps directly relevant to clinical adoption: both discrimination and calibration slopes were lower for patients of Hispanic or Latino ethnicity (AUROC 0.64, slope 0.48), self-pay patients (0.58, 0.42), and the smaller other-race stratum (0.56, 0.29); for the latter two strata the AUROC confidence intervals include 0.5. In all three strata the model over-predicted absolute event rates (observed vs predicted 30-day any-revisit: 13.5% vs 19.7%, other race; 15.0% vs 21.0%, Hispanic or Latino; 16.3% vs 20.1%, self-pay), mirroring the cohort-wide over-prediction (calibration-in-the-large −0.84 to −0.42). Discrimination near chance cannot be remedied by recalibration alone, and the affected strata are among the smallest in the cohort (n = 156–400), consistent with under-representation as a contributor. Targeted data enrichment or model revision, rather than post-hoc adjustment alone, would be required before clinical use, and subgroup audits of this kind belong in any foundation-model evaluation intended to support adoption decisions. Readiness is also version-specific: foundation models within vendor ecosystems are updated on the vendor’s schedule, so this evaluation attaches to the model version tested, and clinical use would require ongoing performance surveillance, with re-evaluation of overall and subgroup performance when the model or the underlying coding practices change.

### Limitations

This study has several limitations. First, the cohort was drawn from a single clinical presentation and a single foundation model, and the evaluation cohort comes from the same network on which Curiosity was pretrained. Although these patients were excluded from training, performance reflects in-distribution coding practices and may not transfer to other conditions, systems, or EHR environments. External validation is required before generalizability can be assumed. Second, Epic Cosmos captures only within-network post-discharge care, so observed revisit rates may be biased downward. Third, the XGBoost baseline used the feature representation from the original Curiosity development paper; although we extended training to the learning-curve plateau and tuned hyperparameters per endpoint, we tested only one family of supervised models, and other well-tuned approaches might narrow or close the discrimination gap. Fourth, subgroup analyses used the any-revisit endpoint rather than the primary admit-revisit endpoint because of limited events per stratum. Subgroup analyses were performed for Curiosity only, so we cannot distinguish model-specific behavior from under-representation of these groups in the source data. Fifth, Curiosity’s predicted probabilities are proportions of 50 simulations, limiting resolution to 2-percentage-point increments and pinning estimates at 0 or 1; this granularity ceiling may modestly attenuate calibration slopes and is most consequential for low-prevalence outcomes such as admit-revisit. Sixth, Curiosity uses only discrete coded events, omitting vital signs, notes, imaging, and medication-administration records that are central to abdominal-pain disposition. Seventh, outcomes were terminal: classification was fixed by the first ED revisit, so a discharge-revisit followed by an admission within 30 days counts only as a discharge-revisit. This does not affect the model comparison but understates admission risk for real-world use. Finally, some outpatient-contact signal reflects system behavior (auto-scheduled follow-up and discharge-initiated telehealth or messaging) rather than patient care-seeking.

## Conclusion

A generative medical event foundation model can represent post-discharge risk as a trajectory rather than a single probability, capturing the timing, sequence, and severity of downstream care for adults discharged after ED evaluation for abdominal pain, and identifying patients most likely to return requiring admission. Subgroup performance gaps must be resolved before clinical use, and whether these capabilities translate into improvements in resource allocation, disposition decisions, discharge counseling, or resource planning remains a key question for prospective study.

## Supporting information

Tripod AI Checklist

## Author contributions

Kent A. McCann (Conceptualization, Data curation, Formal analysis, Investigation, Methodology, Software, Writing—original draft, Writing—review & editing), Donald S. Wright (Conceptualization, Investigation, Resources, Writing—review & editing), Mark S. Iscoe (Investigation, Writing—original draft, Writing—review & editing), Edward R. Melnick (Investigation, Supervision, Writing—review & editing), Lucila Ohno-Machado (Investigation, Supervision, Writing—review & editing), Daniella Meeker (Formal analysis, Investigation, Resources, Writing—review & editing), Arjun K. Venkatesh (Conceptualization, Investigation, Supervision, Writing—review & editing), Rohit B. Sangal (Conceptualization, Investigation, Supervision, Writing—original draft, Writing— review & editing), and Andrew J. Loza (Conceptualization, Formal analysis, Investigation, Methodology, Supervision, Writing—original draft, Writing—review & editing). Drs. Sangal and Loza contributed equally as co-senior authors. Dr. McCann had full access to all the data in the study and takes responsibility for the integrity of the data and the accuracy of the data analysis.

## Data availability

Deidentified electronic health record data underlying these analyses reside in Epic Cosmos and are available to investigators with approved access through Epic Systems. The Curiosity model used in this study is described in Waxler et al and is hosted within the Epic Cosmos environment. Analytic code resides within Epic Cosmos and is available to investigators with approved Cosmos access.

## Use of artificial intelligence

Claude Opus (versions 4.6 – 5.0; Anthropic) and Claude Fable (Version 5; Anthropic) were used to assist in drafting and revising the analytic pipeline and manuscript. The authors take full responsibility for the integrity of the analysis and the content of the manuscript.

