## Supplemental Materials for "Evaluation of a Generative Medical Event Foundation Model for Predicting Post-Discharge Trajectories in Emergency Department Abdominal Pain"

**Supplementary Appendix**

**Contents**

Supplementary Methods
Figure S1. Convergence of Trajectory Accuracy and Primary-Endpoint Discrimination by Simulation Count
Figure S2. XGBoost Baseline Learning Curve
Table S1. Encounter Type Grouping and Per-Timeline Frequencies
Table S2. Trajectory State Space
Table S3. XGBoost Baseline Hyperparameters
Table S4. Individual-Level Calibration by Outcome and Horizon
Table S5. Characteristics of the XGBoost Training and Evaluation Cohorts

**Supplementary Methods**

**Bootstrap confidence intervals for per-outcome discrimination.** For each of the nine outcome-by-horizon combinations, 95% confidence intervals for AUROC and AUC-PR were constructed using 1,000 bias-corrected and accelerated (BCa) bootstrap iterations, with patient as the resampling unit to preserve within-patient correlation between Curiosity and XGBoost predictions. AUC-PR was reported without a formal paired significance test.

**Trajectory edit distance.** Edit distance quantifies the number of trajectory elements that must be added, removed, or substituted to transform one predicted trajectory into the observed one.[1] McNemar’s tests for the most-probable, three-most-probable, and five-most-probable match rates used Yates continuity correction.[2]

**Hierarchical bootstrap for stepwise calibration.** To account for uncertainty at both the patient and simulation levels, we used a two-level hierarchical bootstrap (1,000 iterations): at each iteration, patients were resampled with replacement, and within each resampled patient, their 50 simulations were also resampled with replacement. This generated a distribution of simulated conditional probabilities for each of the 45 transitions.[3] The 95% CI reported in the main text is the 2.5th to 97.5th percentile range of the median absolute simulated-versus-observed difference across all 45 transitions.

**Individual-level calibration.** Curiosity’s predicted probabilities are proportions of 50 simulations. Before logit transformation, proportions were mapped by the empirical-logit continuity correction $p=(k+0.5)/51$, where $k$ is the number of simulations containing the outcome, the standard transform for proportions estimated from a finite number of draws. Calibration slope was estimated by free-slope logistic regression of the observed outcome on the transformed logit; calibration-in-the-large is the intercept of the corresponding model with the slope fixed at 1. The integrated calibration index was computed on the probability scale. Subgroup calibration slopes (Table 3) used the same estimator. As a sensitivity analysis, raw proportions clamped to $[{10}^{-6},1-{10}^{-6}]$ without continuity correction yielded systematically attenuated slopes (0.20–0.77 across the nine endpoints, vs 0.72–1.11 under the empirical logit), because the large mass of patients with zero positive simulations maps to an extreme logit ($\approx-13.8$); the direction and approximate magnitude of calibration-in-the-large were unchanged. The 0.02 quantization of simulation-based probabilities may still modestly attenuate estimated slopes.

**XGB calibration.** Calibration metrics for XGBoost were computed identically, except that slope and calibration-in-the-large were estimated by standard logistic recalibration of logit-transformed probabilities; the empirical-logit adjustment was unnecessary because XGBoost probabilities are continuous rather than simulation proportions.

**Choice of simulation count.** Because Curiosity’s predicted probabilities are proportions of a finite number of simulations, metrics computed from them carry a finite-sample bias whose direction is set by the metric’s curvature; for concave metrics such as AUROC this bias is downward, diminishes as the simulation count rises, and can be modeled to predict performance gains from increased simulation counts.[4] To assess the magnitude of potential performance benefits from increased sample size, we subsampled N of each patient’s 50 simulations without replacement (N = 5 to 50, 200 resamples per N) and recomputed 30-day admit-revisit AUROC. Mean AUROC rose monotonically with N, reaching the reported value of 0.832 at 50 simulations and projected approximately 0.01 additional AUROC at 100 simulations (0.844, 95% CI 0.829–0.863).

**Software versions.** Python 3.13; pandas 2.2.3, numpy 2.1.3, scipy 1.15.3, statsmodels 0.14.5, scikit-learn 1.6.1, xgboost 3.1.1, optuna 3.5.0. Figures were generated in matplotlib 3.10.0 and plotly 5.19.0; final panel layout was assembled in Figma.

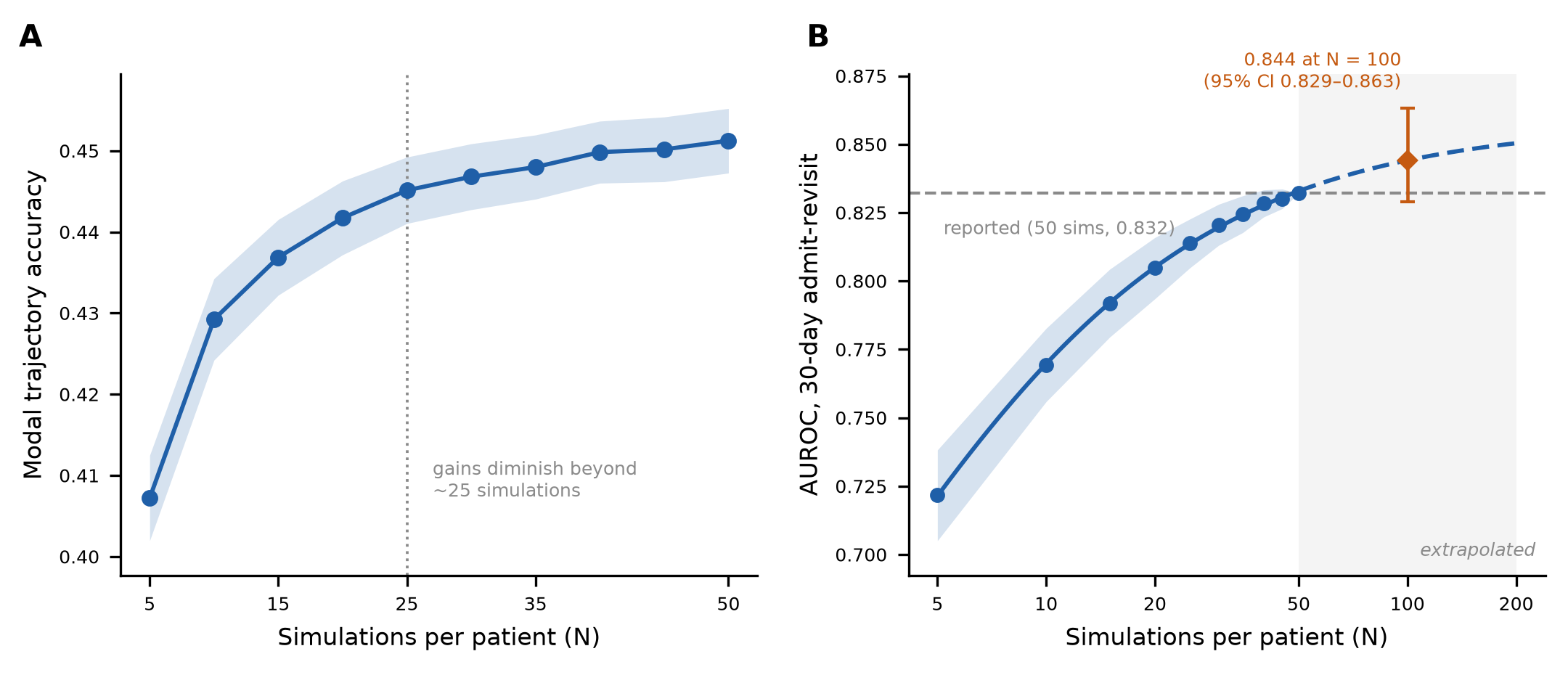

**Figure S1.** Convergence of Trajectory Accuracy and Primary-Endpoint Discrimination by Simulation Count. (A) Modal trajectory classification accuracy (mean ± SD across 200 bootstrap resamples at each simulation count, N = 5 to 50) for the 3,000-patient evaluation cohort. Accuracy improvements diminished beyond approximately 25 simulations per patient, supporting the selection of 50 simulations for trajectory-level analyses. (B) AUROC for the primary endpoint (30-day admit-revisit) computed from subsamples of N of each patient’s 50 simulations (without replacement; mean ± SD across 200 resamples per N), with the finite-sample bias-decay curve fitted to the observed sweep (solid line) and extrapolated beyond N = 50 (dashed line, shaded region).[4] The reported AUROC of 0.832 at 50 simulations is projected to rise to 0.844 (95% CI 0.829–0.863) at N = 100 (orange diamond; parametric bootstrap CI).

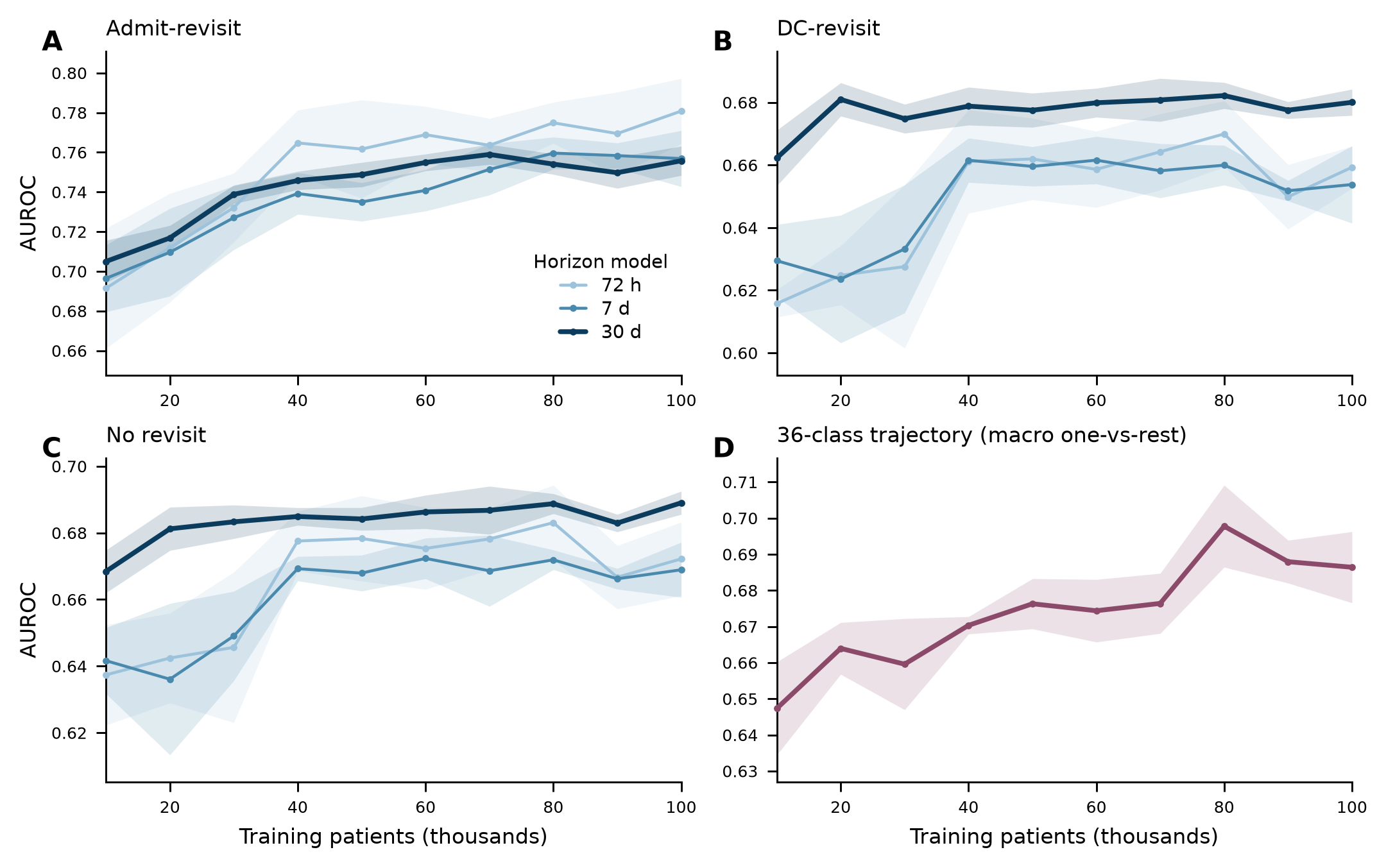

**Figure S2. XGBoost Baseline Learning Curve.** Discrimination of the XGBoost baseline as a function of training-set size (10,000 to 100,000 patients; 5 random seeds per size; mean $\pm$ SD). For the primary endpoint (30-day admit-revisit), AUROC rose from 0.71 at 10,000 training patients and plateaued by approximately 60,000 patients (0.75–0.76), changing by <0.01 per doubling thereafter. The 36-class trajectory model rose through approximately 80,000 patients, with mean macro one-vs-rest AUROC within 0.012 across the 80,000–100,000 range. Each of the four models was subsequently hyperparameter-tuned at 100,000 patients (Table S3); the tuned models served as the comparator for all head-to-head analyses (Table 2).

**Table S1. Encounter Type Grouping and Per-Timeline Frequencies.** Distinct encounter types observed in the evaluation cohort were consolidated into the higher-level categories used for trajectory classification (In-Person, Tele/Msg, Lab/Imaging, Pharmacy, ED Visit, Admission). Columns report the per-category counts per patient timeline in the simulated and observed cohorts. Of all encounters within the 30-day window, 83.7% of observed (8,474/10,130) and 85.4% of simulated (431,717/505,501) encounters mapped to one of the categories listed above and were included in trajectory construction; encounters not mapping to any Grouped Category were excluded. Encounter types with simulated frequency $<$ 0.001 are omitted for space.

| **Encounter Type** | **Grouped** **Category** | **Sim. freq** | **Obs. freq** |
| --- | --- | --- | --- |
| Telephone | Telehealth/Message | 0.734 | 0.740 |
| Office_Visit | In-Person Visit | 0.456 | 0.509 |
| Patient_Message | Telehealth/Message | 0.283 | 0.289 |
| Emergency | DC-Revisit | 0.281 | 0.259 |
| Orders_Only | Telehealth/Message | 0.277 | 0.299 |
| Hospital_Outpatient_Visit | In-Person Visit | 0.241 | 0.249 |
| Refill | Pharmacy | 0.206 | 0.217 |
| Pharmacy_Visit | Pharmacy | 0.102 | 0.097 |
| Patient_Outreach | Telehealth/Message | 0.073 | 0.076 |
| Lab | Lab/Imaging | 0.072 | 0.068 |
| Ancillary_Orders | Telehealth/Message | 0.069 | 0.057 |
| Documentation |  | 0.058 | 0.061 |
| Anesthesia |  | 0.056 | 0.065 |
| Home_Care_Visit | In-Person Visit | 0.056 | 0.044 |
| Emergency_to_Inpatient | Admit-Revisit | 0.052 | 0.056 |
| Abstract |  | 0.052 | 0.065 |
| Surgery |  | 0.051 | 0.062 |
| Telemedicine | Telehealth/Message | 0.046 | 0.035 |
| MyChart | Telehealth/Message | 0.045 | 0.045 |
| Other_Hospital_Encounters | In-Person Visit | 0.042 | 0.055 |
| Appointment | In-Person Visit | 0.033 | 0.027 |
| Ancillary_Procedure |  | 0.031 | 0.040 |
| Nurse_Triage |  | 0.029 | 0.028 |
| Clinical_Support | In-Person Visit | 0.026 | 0.069 |
| Transcribe_Orders |  | 0.025 | 0.021 |
| External_Contact |  | 0.024 | 0.023 |
| Clinical_Documentation_Only |  | 0.021 | 0.013 |
| Home_Care_Update | Telehealth/Message | 0.021 | 0.022 |
| Routine_Prenatal | In-Person Visit | 0.015 | 0.019 |
| Infusion |  | 0.015 | 0.023 |
| Inpatient_Admission | Admit-Revisit | 0.015 | 0.013 |
| Prep_for_Procedure |  | 0.014 | 0.014 |
| Procedure_visit | In-Person Visit | 0.012 | 0.011 |
| Lab_Requisition | Lab/Imaging | 0.011 | 0.006 |
| Consult | In-Person Visit | 0.009 | 0.004 |
| Follow-Up | In-Person Visit | 0.009 | 0.011 |
| E-Visit | Telehealth/Message | 0.009 | 0.007 |
| Nurse_Only |  | 0.008 | 0.009 |
| Case_Management |  | 0.007 | 0.008 |
| Anticoagulation_Visit |  | 0.006 | 0.007 |
| Treatment |  | 0.006 | 0.008 |
| Plan_of_Care_Documentation |  | 0.005 | 0.007 |
| Initial_Prenatal | In-Person Visit | 0.005 | 0.005 |
| Pre-Admission_Testing |  | 0.004 | 0.003 |
| Immunization |  | 0.004 | 0.005 |
| Allied_Health |  | 0.004 | 0.004 |
| Radiology_Appointment | Lab/Imaging | 0.004 | 0.001 |
| Walk-In | In-Person Visit | 0.003 | 0.002 |
| Social_Work |  | 0.003 | 0.001 |
| Community_Orders |  | 0.003 | 0.005 |
| Evaluation |  | 0.002 | 0.001 |
| Education |  | 0.002 | 0.001 |
| E-Consult |  | 0.002 | 0.003 |
| Nutrition |  | 0.001 | 0.001 |
| Hospital_OP_Visit_to_Inpatient | Admit-Revisit | 0.001 |  |
| Hospital | Admit-Revisit | 0.001 | 0.001 |
| Hospice_Admission | In-Person Visit | 0.001 | 0.001 |
| Urgent_Care | In-Person Visit | 0.001 | 0.001 |

**Table S2. Trajectory State Space.** Each row represents one of the 36 possible post-discharge trajectory classes, defined by the combination of outpatient contact (present or absent) and clinical outcome (no ED revisit, DC-revisit, or admit-revisit) at each time window. ED revisit outcomes are terminal: patients who revisit the ED exit subsequent time windows (blank cells). The Length column indicates the number of trajectory elements in that class (12-token vocabulary). DC-revisit = ED revisit ending in discharge; admit-revisit = ED revisit ending in admission. OP = outpatient contact; “—” = no outpatient contact during that window. Bold text marks the terminal outcome for each trajectory class.

| **Class** | **Len** | **0–72h OP** | **0–72h Outcome** | **72h–7d OP** | **72h–7d Outcome** | **7–30d OP** | **7–30d Outcome** |
| --- | --- | --- | --- | --- | --- | --- | --- |
| 1 | 1 |  | **DC-revisit** |  |  |  |  |
| 2 | 1 |  | **Admit-revisit** |  |  |  |  |
| 3 | 2 | OP | **DC-revisit** |  |  |  |  |
| 4 | 2 | OP | **Admit-revisit** |  |  |  |  |
| *Trajectories terminating in the 72h–7d window* | | | | | | | |
| 5 | 2 |  | No revisit |  | **DC-revisit** |  |  |
| 6 | 2 |  | No revisit |  | **Admit-revisit** |  |  |
| 7 | 3 |  | No revisit | OP | **DC-revisit** |  |  |
| 8 | 3 |  | No revisit | OP | **Admit-revisit** |  |  |
| 9 | 3 | OP | No revisit |  | **DC-revisit** |  |  |
| 10 | 3 | OP | No revisit |  | **Admit-revisit** |  |  |
| 11 | 4 | OP | No revisit | OP | **DC-revisit** |  |  |
| 12 | 4 | OP | No revisit | OP | **Admit-revisit** |  |  |
| *Trajectories reaching the 7–30 day window* | | | | | | | |
| 13 | 3 |  | No revisit |  | No revisit |  | **No revisit** |
| 14 | 3 |  | No revisit |  | No revisit |  | **DC-revisit** |
| 15 | 3 |  | No revisit |  | No revisit |  | **Admit-revisit** |
| 16 | 4 |  | No revisit |  | No revisit | OP | **No revisit** |
| 17 | 4 |  | No revisit |  | No revisit | OP | **DC-revisit** |
| 18 | 4 |  | No revisit |  | No revisit | OP | **Admit-revisit** |
| 19 | 4 |  | No revisit | OP | No revisit |  | **No revisit** |
| 20 | 4 |  | No revisit | OP | No revisit |  | **DC-revisit** |
| 21 | 4 |  | No revisit | OP | No revisit |  | **Admit-revisit** |
| 22 | 5 |  | No revisit | OP | No revisit | OP | **No revisit** |
| 23 | 5 |  | No revisit | OP | No revisit | OP | **DC-revisit** |
| 24 | 5 |  | No revisit | OP | No revisit | OP | **Admit-revisit** |
| 25 | 4 | OP | No revisit |  | No revisit |  | **No revisit** |
| 26 | 4 | OP | No revisit |  | No revisit |  | **DC-revisit** |
| 27 | 4 | OP | No revisit |  | No revisit |  | **Admit-revisit** |
| 28 | 5 | OP | No revisit |  | No revisit | OP | **No revisit** |
| 29 | 5 | OP | No revisit |  | No revisit | OP | **DC-revisit** |
| 30 | 5 | OP | No revisit |  | No revisit | OP | **Admit-revisit** |
| 31 | 5 | OP | No revisit | OP | No revisit |  | **No revisit** |
| 32 | 5 | OP | No revisit | OP | No revisit |  | **DC-revisit** |
| 33 | 5 | OP | No revisit | OP | No revisit |  | **Admit-revisit** |
| 34 | 6 | OP | No revisit | OP | No revisit | OP | **No revisit** |
| 35 | 6 | OP | No revisit | OP | No revisit | OP | **DC-revisit** |
| 36 | 6 | OP | No revisit | OP | No revisit | OP | **Admit-revisit** |

**Table S3. XGBoost Baseline Hyperparameters.** Panel A: fixed hyperparameters following Waxler et al,[5] used for the learning-curve sweep (Figure S2). Separate three-outcome models (no ED revisit, DC-revisit, admit-revisit) were trained for each time horizon, and a fourth model predicted the trajectory class (out of 36 classes). Panel B: Bayesian hyperparameter optimization (Optuna) applied independently to each of the four models at the 100,000-patient plateau; the tuned, seed-ensembled models served as the comparator for all head-to-head analyses. The search was run on a stratified 50,000-patient subsample with final models refit on the full training sample. No class-imbalance resampling or reweighting was applied.

| *Panel A. Fixed development hyperparameters (learning-curve sweep)* |  |
| --- | --- |
| **Parameter** | **Value** |
| Number of estimators | 10,000 |
| Maximum depth | 6 |
| Learning rate | 0.1 |
| Subsample fraction | 0.8 |
| Early stopping | 100 |
| Validation split | Stratified 10% held-out |
| *Panel B. Hyperparameter tuning at the 100,000-patient plateau (final comparator)* |  |
| **Parameter** | **Search range** |
| Maximum depth | 3–10 |
| Learning rate | 0.01–0.3 (log) |
| Subsample fraction | 0.5–1.0 |
| Column subsample per tree | 0.4–1.0 |
| Minimum child weight | 1–20 (log) |
| Gamma | ${10}^{-8}$–5 (log) |
| L1 regularization (alpha) | ${10}^{-8}$–10 (log) |
| L2 regularization (lambda) | ${10}^{-8}$–10 (log) |
| Trials per endpoint | 40 (Optuna, 3-fold cross-validation) |
| Early stopping / estimator ceiling | 50 rounds / 3,000 |
| Final refit | Full 100,000-patient sample; 5-seed ensemble |
| Selected values per endpoint | Best-trial values (parameter order: maximum depth, learning rate, subsample, column subsample, minimum child weight, gamma, L1, L2):  72h – 6, 0.010, 0.67, 0.52, 9.2, 1.2e-05, 1.3e-08, 4.9e-06  7d – 8, 0.011, 0.66, 0.62, 19.2, 7.2e-06, 6.1e-03, 3.1e-06 30d – 7, 0.010, 0.75, 0.60, 9.6, 6.6e-04, 2.3e-04, 2.6e-08 trajectory – 6, 0.010, 0.52, 0.66, 7.6, 6.5e-07, 1.4e-05, 6.5 |

**Table S4. Individual-Level Calibration by Outcome and Horizon.** Logistic calibration slope (1.0 indicates ideal calibration; values below 1 indicate over-extreme probabilities), calibration-in-the-large (CITL; 0 indicates ideal, negative values indicate over-prediction on average), and integrated calibration index (ICI; mean absolute difference between predicted and lowess-smoothed observed probability, 0 indicates ideal) for Curiosity and the tuned XGBoost models across the nine outcome-by-horizon endpoints in the 3,000-patient evaluation cohort. XGBoost slope and CITL were estimated on logit-transformed probabilities without the empirical-logit adjustment (see Supplementary Methods). Slope and CITL were estimated on empirical-logit-transformed simulation proportions, $p=(k+0.5)/51$ (Supplementary Methods); ICI is computed on the probability scale. Confidence intervals are percentile bootstrap intervals (1,000 resamples). A sensitivity analysis using $\epsilon$-clamped raw proportions in place of the empirical-logit transform yielded attenuated slopes (0.20–0.77; Supplementary Methods). ICI scales with outcome prevalence and is therefore largest for any revisit at 30 days.

| **Outcome** | **Horizon** | **Calibration slope (95% CI)** | **CITL (95% CI)** | **ICI (95% CI)** |
| --- | --- | --- | --- | --- |
| Any revisit | 72h | 0.77 (0.65, 0.91) | −0.62 (−0.80, −0.44) | 0.030 (0.023, 0.037) |
|  | 7d | 0.76 (0.63, 0.90) | −0.55 (−0.71, −0.41) | 0.039 (0.030, 0.049) |
|  | 30d | 0.82 (0.71, 0.93) | −0.43 (−0.54, −0.32) | 0.051 (0.040, 0.063) |
| DC-revisit | 72h | 0.72 (0.55, 0.91) | −0.65 (−0.87, −0.48) | 0.027 (0.021, 0.033) |
|  | 7d | 0.80 (0.65, 0.96) | −0.57 (−0.75, −0.41) | 0.035 (0.027, 0.043) |
|  | 30d | 0.83 (0.71, 0.96) | −0.44 (−0.55, −0.32) | 0.045 (0.035, 0.057) |
| Admit-revisit | 72h | 1.11 (0.94, 1.33) | −0.84 (−1.19, −0.54) | 0.007 (0.005, 0.012) |
|  | 7d | 1.06 (0.90, 1.27) | −0.68 (−0.96, −0.41) | 0.012 (0.008, 0.016) |
|  | 30d | 1.03 (0.89, 1.23) | −0.42 (−0.63, −0.22) | 0.015 (0.010, 0.022) |
| XGBoost: Any revisit | 72h | 1.18 (0.97, 1.40) | −0.03 (−0.21, 0.13) | 0.008 (0.006, 0.015) |
|  | 7d | 1.15 (0.94, 1.35) | −0.07 (−0.20, 0.06) | 0.010 (0.008, 0.019) |
|  | 30d | 1.10 (0.97, 1.24) | −0.04 (−0.14, 0.05) | 0.010 (0.007, 0.023) |
| XGBoost: DC-revisit | 72h | 1.10 (0.81, 1.35) | −0.06 (−0.25, 0.12) | 0.005 (0.005, 0.012) |
|  | 7d | 1.11 (0.87, 1.32) | −0.06 (−0.23, 0.08) | 0.008 (0.007, 0.015) |
|  | 30d | 1.06 (0.91, 1.20) | −0.01 (−0.13, 0.10) | 0.011 (0.009, 0.021) |
| XGBoost: Admit-revisit | 72h | 1.31 (1.05, 1.58) | 0.03 (−0.29, 0.31) | 0.006 (0.004, 0.010) |
|  | 7d | 1.26 (1.00, 1.51) | −0.08 (−0.34, 0.13) | 0.005 (0.004, 0.010) |
|  | 30d | 1.17 (0.99, 1.39) | −0.09 (−0.28, 0.09) | 0.006 (0.005, 0.014) |

**Table S5. Characteristics of the XGBoost Training and Evaluation Cohorts.** Baseline characteristics of the XGBoost training cohort (drawn from the Epic Cosmos training partition using the same 2022 index-encounter eligibility criteria) and the 3,000-patient evaluation cohort (drawn from the non-training partition; Table 1). The cohorts do not overlap. Descriptive statistics are presented as median [IQR] for continuous variables and n (%) for categorical variables. The 30-day outcome is the terminal outcome classification used throughout (no ED revisit, ED revisit ending in discharge [DC-revisit], or ED revisit ending in admission [admit-revisit]); demographic and outcome distributions were closely similar across cohorts. Emergency Severity Index (ESI) levels are collapsed (1–2 and 4–5) in accordance with the Epic Cosmos minimum cell-size policy. The private, other, or unspecified insurance row combines private and miscellaneous or unspecified coverage categories to permit alignment across cohorts.

|  | **Training cohort** | **Evaluation cohort** |
| --- | --- | --- |
| n | 100,000 | 3,000 |
| Age, median [IQR] | 47.0 [36.0–61.0] | 47.0 [36.0–60.3] |
| **Revisits (cumulative)** |  |  |
| 72 Hour | 5,997 (6.0%) | 162 (5.4%) |
| 7 Day | 9,380 (9.4%) | 249 (8.3%) |
| 30 Day | 18,023 (18.0%) | 506 (16.9%) |
| **30-Day Outcome** |  |  |
| No ED revisit | 81,997 (82.0%) | 2,494 (83.1%) |
| DC-revisit | 13,197 (13.2%) | 383 (12.8%) |
| Admit-revisit | 4,826 (4.8%) | 123 (4.1%) |
| **Sex** |  |  |
| Female | 64,173 (64.1%) | 1,958 (65.3%) |
| Male | 34,996 (35.0%) | 1,019 (34.0%) |
| Other/Unspecified | 836 (0.8%) | 23 (0.8%) |
| **Race** |  |  |
| White | 57,028 (57.0%) | 1,738 (57.9%) |
| Black or African American | 20,686 (20.7%) | 627 (20.9%) |
| Not Reported | 12,084(12.1%) | 348 (11.6%) |
| Other | 10,202 (10.2%) | 287 (9.6%) |
| **Ethnicity** |  |  |
| Not Hispanic or Latino | 71,234 (71.2%) | 2,153 (71.8%) |
| Not Reported | 14,972 (15.0%) | 447 (14.9%) |
| Hispanic or Latino | 13,794 (13.8%) | 400 (13.3%) |
| **Acuity Level** |  |  |
| ESI 1–2 – Immediate/Emergent | 6,465 (6.5%) | 193 (6.4%) |
| ESI 3 – Urgent | 89,179 (89.2%) | 2,672 (89.1%) |
| ESI 4–5 – Less/Non-Urgent | 3,640 (3.6%) | 111 (3.7%) |
| Unspecified | 716 (0.7%) | 24 (0.8%) |
| **Insurance** |  |  |
| Private, other, or unspecified | 65,589 (65.6%) | 1,952 (65.1%) |
| Medicaid | 14,685 (14.7%) | 443 (14.8%) |
| Medicare | 13,258 (13.3%) | 402 (13.4%) |
| Self-pay | 6,468 (6.5%) | 203 (6.8%) |
