## Supplementary material for "Evaluation of a Generative Medical Event Foundation Model for Predicting Post-Discharge Trajectories in Emergency Department Abdominal Pain": Tripod AI Checklist

**TRIPOD+AI Checklist**
Post-ED Trajectory Prediction in Abdominal Pain with a Generative Medical Event Model

*Checklist from Collins GS, Moons KGM, Dhiman P, et al. TRIPOD+AI statement: updated guidance for reporting clinical prediction models that use regression or machine learning methods. BMJ. 2024;385:e078378. D = development; E = evaluation. This study evaluates an existing model (Curiosity) and develops a comparator (XGBoost).*

| **Item** | **Checklist item (abbreviated)** | **Location / response** |
| --- | --- | --- |
| **Title and Abstract** | | |
| 1 | Identify the study as developing or evaluating a prediction model, the target population, and the outcome | Title: population (abdominal pain discharged from ED), outcome (post-ED trajectory), and model type are identified |
| 2 | Abstract addressing TRIPOD+AI for Abstracts items | Structured abstract: objective, data source, eligibility, sample size, outcomes, comparator, results with CIs, conclusion |
| **Introduction** | | |
| 3a | Healthcare context (prognostic) and rationale, with references to existing models | Introduction: Background and Importance; existing revisit models referenced |
| 3b | Target population, intended purpose in care pathway, intended users | Introduction: Importance and Goals; intended use at ED discharge by clinicians; elaborated in Discussion (Beyond Binary Prediction) |
| 3c | Known health inequalities between sociodemographic groups | Introduction: Background (variation in revisit risk and post-discharge outcomes across demographic and socioeconomic groups) |
| 4 | Study objectives: development, evaluation, or both | Introduction: Goals of This Investigation (evaluation of Curiosity against a task-specific XGBoost comparator developed for this study) |
| **Methods** | | |
| 5a | Sources of data for development and evaluation datasets; rationale; representativeness | Methods: Study Design and Setting (Epic Cosmos network EHR, >300 million patients); Comparator (training cohorts drawn from the Cosmos training partition; final comparator trained on 100,000 patients) |
| 5b | Dates of participant data, accrual, end of follow-up | Methods: Selection of Participants (index ED encounters Jan 1–Dec 31, 2022; 30-day follow-up); Comparator (training cohorts identified with the same 2022 index-encounter criteria) |
| 6a | Study setting, number and location of centres | Methods: Study Design and Setting (>300 health systems; United States, Canada, Middle East) |
| 6b | Eligibility criteria | Methods: Selection of Participants (age ≥18, ≥2 encounters, structured chief complaint coded as abdominal pain, discharged; first qualifying encounter retained) |
| 6c | Treatments received and their handling | Not applicable: treatments during the index visit were not used as predictors or outcomes; post-discharge medication orders contribute to outpatient-contact categories (Table S1) |
| 7 | Data pre-processing and quality checking, including across sociodemographic groups | Methods: Outcomes and Table S1 (encounter-type mapping; unmapped encounters excluded); no imputation. Pre-processing identical for all patients |
| 8a | Outcome definition, time horizon, assessment, consistency across groups | Methods: Outcomes (DC-revisit, admit-revisit, outpatient contact; 72-hour/7-day/30-day windows; terminal revisit convention; observation vs inpatient handling). Derived identically from structured encounter types for all patients |
| 8b | Qualifications of outcome assessors (if subjective) | Not applicable: outcomes derived programmatically from structured encounter types |
| 8c | Blinding of outcome assessment | Not applicable: outcomes computed programmatically, independent of model predictions |
| 9a | Choice of initial predictors and pre-selection (D) | Methods: Comparator (event counts over the same feature vocabulary used as Curiosity input; no further pre-selection) |
| 9b | Definition, timing, and measurement of predictors | Methods: Trajectory Simulation (most recent 8,192 events before discharge for Curiosity) and Comparator (complete pre-index history for XGBoost); all predictors measured before the prediction time point |
| 9c | Qualifications of predictor assessors (if subjective) | Not applicable: predictors are recorded EHR events |
| 10 | Sample size justification for development and evaluation | Methods: Selection of Participants (evaluation cohort of 3,000 chosen to balance the computational cost of 50 simulations per patient against the precision of the primary admit-revisit comparison) and Comparator (training size extended along a 10,000–100,000-patient learning curve to its plateau; final models trained at 100,000, so the comparison is not limited by training-set size; Figure S2). Achieved precision for the primary comparison is reflected in the Table 2 bootstrap confidence intervals (123 events; AUROC 95% CI half-width ∼0.04) |
| 11 | Missing data handling | Methods: Selection of Participants (inputs derived directly from recorded events; no imputation); missingness of structured fields reported in Table 1 (not-reported categories) |
| 12a | Data use and partitioning; leakage | Methods: Study Design and Setting, Selection of Participants, Comparator (evaluation cohort from non-training partition; XGBoost trained on separate training-partition cohorts; no patient overlap) |
| 12b | Predictor handling (functional form, transformation) | Methods: Comparator (counts of clinical events); Trajectory Simulation (event sequences; tokenization per Waxler et al) |
| 12c | Model type, building steps, hyperparameter tuning, internal validation (D) | Methods: Comparator and Table S3 (learning-curve sweep 10,000–100,000 patients at fixed development hyperparameters, 5 seeds per size; per-endpoint Bayesian hyperparameter tuning at 100,000 patients, 40 Optuna trials with 3-fold cross-validation; final 5-seed ensembles; early stopping on a stratified validation split); Curiosity used pretrained with default settings, no fine-tuning |
| 12d | Heterogeneity across clusters | Not examined: performance was not stratified by health system (de-identified data; system identifiers not analyzed) |
| 12e | Performance measures and plots, and model comparison methods | Methods: Primary Data Analysis (AUROC, AUC-PR, DeLong tests, trajectory accuracy, edit distance, stepwise calibration, calibration slope/CITL/ICI for both models); Curiosity individual-level calibration estimated on empirical-logit-transformed simulation proportions, XGBoost by standard logistic recalibration of logit-transformed probabilities (Supplementary Methods; Table S4); primary comparison designated |
| 12f | Model updating / recalibration (E) | Methods: Primary Data Analysis (predictions evaluated as generated; no post hoc recalibration) |
| 12g | How model predictions were calculated (E) | Methods: Trajectory Simulation (50 simulations per patient; predicted probability = proportion of simulations containing the outcome) |
| 13 | Class imbalance methods | Methods: Primary Data Analysis (none used; AUC-PR reported against observed base rates) |
| 14 | Approaches to address model fairness | Methods/Results: Subgroup Performance (discrimination and calibration by sex, age, acuity, race, ethnicity, insurance; Table 3) |
| 15 | Model output and any thresholds | Methods: Primary Data Analysis and Results (probabilities from simulation proportions; top-5% groups in Figure 3 are a descriptive display slice, not a decision threshold) |
| 16 | Differences between development and evaluation data | Methods: Study Design and Setting and Comparator (same network; disjoint training/evaluation partitions); Discussion: Limitations (in-distribution evaluation) |
| 17 | Ethics approval | Methods: Study Design and Setting (exempted from human subjects review by the Yale University Institutional Review Board) |
| **Open Science** | | |
| 18a | Funding source and role | Acknowledgments: Funding/Support and Role of the Funder/Sponsor (no external funding) |
| 18b | Conflicts of interest for all authors | Acknowledgments: Conflict of Interest Disclosures (Drs Loza and Meeker are coauthors of the Curiosity development report, Waxler et al; no other conflicts reported) |
| 18c | Protocol availability | No prospective analysis protocol was prepared (stated here) |
| 18d | Registration | The study was not registered (stated here) |
| 18e | Data availability | Acknowledgments: Data Sharing Statement (Epic Cosmos; access through Epic Systems) |
| 18f | Code availability | Acknowledgments: Data Sharing Statement (analytic code available to investigators with approved Epic Cosmos access) |
| **Patient and Public Involvement** | | |
| 19 | Patient and public involvement | Patients and the public were not involved in the design, conduct, reporting, or dissemination of this study (stated here) |
| **Results** | | |
| 20a | Flow of participants, numbers with and without outcome | Results: Study Population (179,310 encounters → 150,030 eligible patients → 3,000 sampled); outcome counts in Table 1 and Table 2 caption |
| 20b | Characteristics of participants, outcome events, missing data | Results: Table 1 (demographics, acuity, cumulative revisits; not-reported categories shown) |
| 20c | Comparison of evaluation vs development data distributions (E) | Supplement: Table S5 (training vs evaluation cohort characteristics; distributions closely similar across age, sex, race, ethnicity, acuity, and insurance) |
| 21 | Numbers of participants and outcome events per analysis | Results: Table 2 caption (base rates: 16.9% any revisit, 12.8% DC-revisit, 4.1% admit-revisit at 30 days); Table 3 (per-stratum events); Simulation Performance (149,720 of 150,000 simulations analyzed) |
| 22 | Full model specification for third-party use (D) | Table S3 (XGBoost development and tuned hyperparameters and feature definition); Curiosity is described in Waxler et al (arXiv:2508.12104) and available within Epic Cosmos (proprietary; access via Epic) |
| 23a | Performance estimates with CIs, including key subgroups | Results: Tables 2 and 3 (bootstrap 95% CIs); Figure 2 (stepwise calibration); Table S4 (calibration metrics with CIs for both models) |
| 23b | Heterogeneity in performance across clusters | Not examined (see item 12d) |
| 24 | Results of model updating (E) | Not applicable: no updating or recalibration performed |
| **Discussion** | | |
| 25 | Overall interpretation including fairness, in context of objectives and prior studies | Discussion: Principal Findings and Beyond Binary Prediction (comparison with prior revisit classifiers and generative-model evaluations; subgroup gaps discussed) |
| 26 | Limitations and their effects on bias, uncertainty, generalizability | Discussion: Limitations (single presentation and model; in-distribution evaluation; within-network capture; single family of supervised comparator models; subgroup endpoint substitution, Curiosity-only subgroup analysis, and fairness gaps; simulation-count granularity ceiling on predicted probabilities; missing input modalities; terminal-outcome classification understating admission risk; system-driven outpatient signal) |
| 27a | Handling of poor quality or unavailable input data at implementation (D) | Discussion: Beyond Binary Prediction (predictions require a longitudinal coded event history mapped to the model’s vocabulary; performance with sparse or fragmented records not evaluated) |
| 27b | Required user interaction and expertise (D) | Partially addressed: Discussion (Beyond Binary Prediction) describes use by clinicians at discharge; no interaction with input data required (predictions derive from recorded EHR events) |
| 27c | Next steps for future research, applicability and generalizability | Discussion: Limitations and closing paragraph (external validation across conditions, systems, and models; prospective implementation research) |
